# A self-report tool for identification of individuals with coronary atherosclerosis – The Swedish CArdioPulmonary BioImage Study

**DOI:** 10.1101/2024.01.24.24301753

**Authors:** Göran Bergström, Eva Hagberg, Elias Björnson, Martin Adiels, Carl Bonander, Ulf Strömberg, Jonas Andersson, Mattias Brunström, Carl-Johan Carlhäll, Gunnar Engström, David Erlinge, Isabel Goncalves, Anders Gummesson, Emil Hagström, Ola Hjelmgren, Stefan James, Magnus Janzon, Lena Jonasson, Lars Lind, Martin Magnusson, Viktor Oskarsson, Johan Sundström, Per Svensson, Stefan Söderberg, Raquel Themudo, Carl Johan Östgren, Tomas Jernberg

## Abstract

**Background:** Coronary atherosclerosis detected by imaging is a marker of elevated cardiovascular risk. However, imaging involves large resources and exposure to radiation. The aim was, therefore, to test whether non-imaging data, specifically data that can be self-reported, could be used to identify individuals with moderate to severe coronary atherosclerosis.

**Methods:** We used data from the population based Swedish CArdioPulmonary BioImage Study (SCAPIS) in individuals with coronary computed tomography angiography (CCTA, n=25,182) and coronary artery calcification score (CACS, n=28,701), aged 50-64 years without previous ischemic heart disease. We developed a risk prediction tool utilizing variables that could be assessed from home (a so-called self-report tool). For comparison, we also developed a tool utilizing variables from laboratory tests, physical examinations and self-report (a so-called clinical tool) and evaluated both models using receiver operating characteristic curve analysis, external validation, and bench-marked against factors in the Pooled Cohort Equation (PCE).

**Results:** The self-report tool (n=14 variables) and the clinical tool (n=23 variables) showed high-to-excellent discriminative ability to identify SIS ≥4 (AUC 0.79 and 0.80, respectively) and significantly better than PCE (AUC 0.76, p<0.001). The tools showed a larger net benefit in clinical decision making at relevant threshold probabilities. The self-report tool identified 65% of all individuals with SIS ≥4 in the top 30% of the highest-risk individuals. Tools developed for CACS ≥100 performed similarly.

**Conclusions:** We have developed a self-report tool which effectively identifies individuals with moderate to severe coronary atherosclerosis. The self-report tool may serve as pre-screening tool towards a cost-effective CT-based screening program for high-risk individuals.

**Clinical Perspective:** *What Is New?:*

- We have developed a self-report tool which with a good-to-excellent discriminative ability identifies individuals with moderate to severe coronary atherosclerosis.
- The self-report tool can be executed from home and has a similar performance to a clinical tool requiring a clinical visit involving blood tests and physical examination.

*What Are the Clinical Implications?:*

- The self-report tool could serve as an initial step towards a cost-effective screening program to identify high-risk individuals or used to identify individuals who would benefit from further risk refinement by cardiac imaging.

## Introduction

Asymptomatic individuals with signs of coronary atherosclerosis on imaging are considered to be at high risk of future ischemic heart disease (IHD).^1–3^ A coronary artery calcification score (CACS) of ≥100, derived from computed tomography (CT) imaging, suggests benefits of statin therapy regardless of LDL concentrations in individuals with an intermediate IHD risk, according to the pooled cohort equation (PCE) risk calculator (7.5-20.0%).^1^ Imaging with coronary CT angiography (CCTA) holds an even bigger promise, since it also visualizes non-calcified coronary atherosclerosis, degree of stenosis, and plaque characteristics; factors that are directly related to an increased risk of future clinical events^4–8^ and that improve risk prediction beyond clinical risk scores such as the PCE.^8–10^ However, the drawbacks of imaging include limited availability, high costs, and risks associated with radiation and use of contrast agents. Therefore, it is of great interest to use non-imaging data to develop tools that identify individuals with high risk of coronary atherosclerosis. These tools could be directly used to identify individuals with an increased risk of IHD or to identify individuals in whom imaging could do more benefit than harm.

To facilitate participation and to reduce costs, it would be advantageous if the tools could be easily administered and based on self-reported data, not requiring a visit to a health care center. A few recent studies show promising results in the self-reported assessment of cardiovascular risk.^11,12^

The aim of this study was to test whether the non-imaging data collected in the Swedish CArdioPulmonary BioImage Study (SCAPIS)^13,14^ could be used to identify individuals with moderate to severe coronary atherosclerosis (defined by the CAD-RADS criteria^15^ as segment involvement score [SIS] ≥4 or CACS ≥100). These cut-off values were selected since previous work with CT^2,16^ and CCTA^7,8,17–19^ have consistently shown that the risk of future IHD is markedly increased at this level of coronary atherosclerosis. The more specific aim was to test if a tool exclusively based on self-reported data could be equally effective as a tool based on the combination of self-reported data and clinical data. The tools were developed and tested in SCAPIS and externally validated in a separate cohort. We included only individuals without previous coronary heart disease and tools were bench-marked against the PCE risk calculator.

## Methods

### The SCAPIS data set

SCAPIS (n=30,154) is a population-based multicenter cohort study of randomly selected men and women aged 50 to 64 years, mainly of European ancestry.^14^ The data collection was performed between 2014 and 2018 at six Swedish University hospitals. A comprehensive examination protocol was used, including cardiac imaging, physical examinations, routine laboratory tests, and questionnaires.^13,14^ SCAPIS was approved by the ethics committee in Umeå, Sweden (number 2010-228-31M), all participants gave written informed consent, and the study protocols adhered to the declaration of Helsinki.

The data set used for external validation (n=1,111) was the single-site SCAPIS pilot trial collected in 2012. This study’s primary objective was to provide insights into the feasibility of performing the nationwide SCAPIS and we therefore used a stratified selection of individuals aimed at low and high socioeconomic areas within the city of Gothenburg.^20^ Participants are unique from the nationwide SCAPIS and different equipment and different staff were used.

### Study populations

Participants without previous coronary heart disease (myocardial infarction or coronary revascularization) and with high-quality imaging of their coronary arteries using CCTA (n=25,182) or high-quality CT imaging for CAC (n=28,701) were selected to address the primary aim of the study (see below for imaging details). Similarly, individuals from the SCAPIS pilot trial (n=872 with CCTA and n=1,062 with CT imaging for CACS) were used for validation.

### Cardiac image acquisition and analyses

As previously described,^14^ we performed cardiac CT scanning with and without contrast agent using a dedicated dual-source CT scanner equipped with a Stellar Detector (Somatom Definition Flash, Siemens Medical Solution, Forchheim, Germany). CCTA were read using the syngo.via software, and the 18-coronary segment model defined by the Society of Cardiovascular Computed Tomography was used to report coronary atherosclerosis as outlined in Supplementary Methods. SIS was calculated as the sum of all coronary artery segments with atherosclerosis.^8^ CACS was analyzed using the method by Agatston.^21^

### Factors used for developing the prediction tools

All factors used to develop the prediction tools are summarized in Supplementary Table S1. Details on how they were collected can be found elsewhere.^13,14^ Information on self-reported health, family history, medication, occupational and environmental exposure, lifestyle, psycho-social well-being, socio-economic status, and other social determinants was collected from questionnaires. Biochemistry analyses were analyzed from venous blood samples collected after an overnight fast. Height, weight, and hip and waist circumference were measured. Physical activity was assessed using an accelerometer. Blood pressure was measured with an automatic device (Omron M10-IT, Omron Health Care Co, Kyoto, Japan). Lung function was measured using spirometry after bronchodilatation. Variables were used in their original scale (not normalized) during model training.

### Outcomes

We used two different outcome variables: SIS ≥4 and CACS ≥100. A detailed rationale for the selected cut-off values is presented in the Supplementary Methods. According to recent consensus by CAD-RADS,^15^ SIS ≥4 and CACS ≥100 represent moderate to severe coronary atherosclerosis.

### Data analysis and statistics

In this study, we developed a self-report tool based on all available self-reported data. Additionally, a clinical tool was developed, using all SCAPIS data, including self-reported data, blood tests, and physical examinations. The purpose of the clinical tool was to serve as a comparative measure to the self-report model. From the SCAPIS baseline data, we identified 105 factors as potential predictors in the self-report tool. A total of 127 factors was identified as potential predictors in the clinical tool (all factors are summarized in Supplementary Table S1). Factors in the self-report tool are possible to report without visiting a healthcare facility. The performance of both of the tools was bench-marked against the 10-year risk of atherosclerotic cardiovascular disease according to the PCE.^1^ We also estimated the average time it took for ten co-authors to fill out the self-report tool when presented in paper format. Descriptive data are presented without imputations, while data used for the prediction tools were imputed using the K-nearest neighbor algorithm with 5 neighbors. For descriptive data, numbers, percentages, mean values (standard deviation), and median values (interquartile range) were calculated. A grid-search was employed in order to optimize model hyperparameters. We followed the TRIPOD statement for transparent reporting.^22^ All analyses were performed using R version 4.1.3.

#### Data reduction

To identify and include the most relevant factors in our tools, we performed data reduction using a combination of data driven techniques (Boruta) and manual techniques (described in detail in the Supplement Methods). In a sensitivity analyses, we also tested a model based on all 127 variables without prior data selection.

#### Development of assessment tools for moderate to severe coronary atherosclerosis

After the data reduction, we used XGBoost (Extreme Gradient Boosting; a decision-tree-based machine learning method) to develop the tools to identify SIS ≥4 and CACS ≥100. The dataset was randomly split into a 75% training set and a 25% test set. The area under the receiver operating characteristic curve (AUC) was calculated and validated in the external study population and compared using the DeLong test.^23^ We avoided overfitting by three strategies. First, hyper-parameter tuning was employed to minimize the risk of overfitting, second, we included an internal-validation subset of the data that the model had never trained on and third, we used a separate cohort as a validation cohort. Our results show that the models appear correctly trained. Calibration curves were constructed and compared between internal and external validation. We also calculated the importance ranking of each variable with XGBoost and the SHapley Additive exPlanations (SHAP)-value. SHAP values represent the relationship between each factor and the outcome for each subject, for which a higher SHAP value confers a higher assessed risk by the XGBoost algorithm.

#### Methodological considerations

Machine-learning methodology was preferred over logistic regression modelling for several reasons. First, the model can account for complex variable relationships with the outcome and interactions between variables without the need for explicit specification of this. Second, model interpretability is improved with possibility of both overall and per-subject variable importance analysis. Third, variable selection procedure is simpler and more accurate.

#### Stratification for age, sex, and socio-economy

Analyses were stratified according to sex and age (50-54, 55-59, and 60-64 years). In addition, to examine the impact of socio-economic factors, analyses were stratified by the individuals’ socioeconomic status based on three different characteristics: education (university degree or lower education), country of birth (born in Sweden or not), and the ability to raise a sum of 20,000 Swedish kronor within a week (equivalent to US$ 1,900).

#### Construction of population-ordered distributions and decision curves

To assess our tools’ performance in a potential screening situation, we constructed population-ordered distribution tables with ten groups of equal size (deciles). We also constructed clinical decision curves to evaluate if our tools gave a net benefit over the PCE at relevant threshold probabilities.^24^

## Results

### Study population

The study inclusion is shown in Supplementary Figure S1. In total, 25,182 individuals were included in the cohort studying SIS ≥4 as an outcome, for whom the characteristics are shown in Table 1. In total, 42.1% of the participants had coronary artery atherosclerosis and 5.1% of women and 19% of men had SIS ≥4. In general, participants with SIS ≥4 had a more severe risk factor profile and their 10-year risk of atherosclerotic cardiovascular disease (according to the PCE) was more than 50% higher than participants with SIS <4 (1.8 times higher in women and 1.5 times in men). In total, 28,701 individuals were included in the cohort that studied CACS ≥100 as an outcome and their characteristics are shown in Supplementary Table S1. In the total population, 37.7% of the participants had positive CACS and 6.1% of women and 22.9% of men had CACS ≥100.

**Table 1:** Clinical characteristics of individuals (n=25,182) with high-quality coronary computed tomography angiography, without known coronary heart disease, stratified by sex and segment involvement score (SIS) ≥4.

|  | Total | Female |  | Male |  |
| --- | --- | --- | --- | --- | --- |
|  | All SIS | SIS < 4 | SIS ≥ 4 | SIS < 4 | SIS ≥ 4 |
| N | 25,182 | 12,094 | 644 | 10,085 | 2359 |
| Sociodemographics |  |  |  |  |  |
| Age, Years | 57.4 ± 4.3 | 57.31 ± 4.3 | 59.62 ± 3.9 | 56.94 ± 4.3 | 59.26 ± 4.1 |
| Education, University degree, n (%) | 11,300 (46.0) | 6010 (49.7) | 242 (37.6) | 4183 (41.5) | 865 (36.7) |
| Born in Sweden - yes, n (%) | 20,893 (83.0) | 10,030 (82.9) | 533 (82.8) | 8455 (83.8) | 1875 (36.7) |
| Can raise 20 000 SEK* within a week - yes, n (%) | 22,515 (89.4) | 10,748 (88.9) | 551 (85.6) | 9152 (90.7) | 2064 (87.5) |
| Anthropometry |  |  |  |  |  |
| Weight at age 20, kg | 66.5 ± 11.3 | 58.0 ± 7.8 | 58.8 ± 9.0 | 73.0 ± 8.9 | 73.6 ± 9.5 |
| Weight, kg | 80.1 ± 15.5 | 72.2 ± 13.0 | 74.6 ± 14.6 | 87.6 ± 13.3 | 89.9 ± 14.5 |
| Height, cm | 173 ± 9.6 | 166 ± 6.4 | 165 ± 6.1 | 180 ± 6.9 | 179 ± 7.0 |
| Waist circumference, cm | 94.0 ± 12.6 | 88.7 ± 11.9 | 93.1 ± 13.2 | 98.6 ± 10.7 | 101.9 ± 11.4 |
| Hip circumference, cm | 102 ± 8.5 | 103 ± 9.7 | 104 ± 10.8 | 102 ± 6.9 | 102 ± 7.4 |
| Smoking status |  |  |  |  |  |
| Current smoker, n (%) | 3215 (12.8) | 1504 (12.4) | 172 (26.7) | 1142 (11.3) | 397 (16.8) |
| Mean pack-years, Years | 7.2 ± 11.7 | 6.8 ± 10.6 | 15.9 ± 15.6 | 6.4 ± 11.5 | 10.5 ± 14.7 |
| Duration of smoking, Years | 11.6 ± 15.2 | 11.8 ± 15.3 | 22.9 ± 17.9 | 9.8 ± 14.6 | 15.1 ± 17.0 |
| Treatment |  |  |  |  |  |
| Cholesterol-lowering medication, n (%) | 1624 (6.4) | 534 (4.4) | 117 (18.2) | 578 (5.7) | 395 (16.7) |
| Antihypertensive medication, n (%) | 4437 (17.6) | 1852 (15.3) | 233 (36.2) | 1554 (15.4) | 798 (33.8) |
| Diabetes medication, n (%) | 717 (2.8) | 198 (1.6) | 49 (7.6) | 273 (2.7) | 197 (8.4) |
| Diagnosis |  |  |  |  |  |
| Diagnosed hypertension, n (%) | 2056 (8.2) | 807 (6.7) | 115 (17.9) | 707 (7.0) | 427 (18.1) |
| Diabetes duration, age (years) | 0.3 (2.6) | 0.2 (2.3) | 1.2 (6.0) | 0.3 (2.1) | 0.9 (4.2) |
| Blood pressure, mmHg |  |  |  |  |  |
| Systolic, mmHg | 126 ± 17 | 122 ± 18 | 130 ± 18 | 128 ± 15 | 133 ± 16 |
| Diastolic, mmHg | 78 ± 11 | 77 ± 11 | 79 ± 11 | 78 ± 10 | 80 ± 10 |
| Clinical chemistry |  |  |  |  |  |
| Total cholesterol, mmol/L | 5.5 ± 1.0 | 5.7 ± 1.0 | 5.8 ± 1.2 | 5.4 ± 1.0 | 5.5 ± 1.1 |
| HDL cholesterol, mmol/L | 1.6 ± 0.5 | 1.9 ± 0.5 | 1.7 ± 0.5 | 1.4 ± 0.4 | 1.4 ± 0.4 |
| LDL, calculated, cholesterol, mmol/L | 3.5 ± 0.9 | 3.4 ± 0.9 | 3.7 ± 1.1 | 3.5 ± 0.9 | 3.6 ± 1.1 |
| Triglycerides, mmol/L | 1.2 ± 0.8 | 1.1 ± 0.6 | 1.4 ± 0.8 | 1.3 ± 0.9 | 1.5 ± 0.9 |
| Glucose, mmol/mL | 5.7 ± 1.0 | 5.5 ± 0.8 | 5.9 ± 1.4 | 5.8 ± 1.0 | 6.2 ± 1.5 |
| HbA1c, mmol/mL | 36.3 ± 5.9 | 35.9 ± 4.9 | 38.6 ± 8.0 | 36.0 ± 5.7 | 38.4 ± 8.8 |
| High-sensitive C- reactive protein, mg/L | 2.1 ± 3.9 | 2.1 ± 3.6 | 2.7 ± 5.2 | 1.9 ± 3.7 | 2.4 ± 5.4 |
| Risk score |  |  |  |  |  |
| Pooled cohort equation, % | 6.2 ± 5.5 | 3.2 ± 2.7 | 5.9 ± 4.3 | 8.4 ± 5.3 | 12.5 ± 6.9 |
| Heredity |  |  |  |  |  |
| Family history of premature myocardial infarction, n (%) | 1619 (6.4) | 821 (6.8) | 79 (12.3) | 515 (5.1) | 204 (8.6) |
| Family history of premature stroke, n (%) | 1468 (5.8) | 780 (6.4) | 56 (8.7) | 469 (4.7) | 163 (6.9) |

\*SEK: Swedish Krona, LDL: low-density lipoprotein, HDL: high-density lipoprotein, HbA1c: glycated hemoglobin

In the validation cohort, a higher proportion of participants born outside Sweden and a lower proportion with university education were observed compared to the SCAPIS dataset. Details on study inclusion and characteristics of the validation cohorts are shown in Supplementary Figure S2, Supplementary Table S3, and Supplementary Table S4.

### Factors selected for developing the self-report and clinical prediction tools

A summary of missing data for each available factor and specification of the relevant factors identified by the Boruta algorithm is shown in Supplementary Table S1. A description of all individual factors can be found at www.scapis.org/portal/variables. Following discussion among lead authors, the self-report tool incorporated 14 factors (Table 2). These factors included seven on demographic and anthropometric information, (sex, age, body weight at age 20, body weight, body height, waist circumference, hip circumference), two on smoking status (cigarette pack-years and smoking duration), four on existing health conditions (lipid lowering medication, antihypertensive medication, diabetes duration and diagnosed hypertension and one on family history (heredity for MI). The clinical tool incorporated 23 factors (Table 2), with all the 14 factors in the self-report model included but also adding vital signs (three factors) and laboratory measurements (six factors). Additional details on data selection can be found in the Supplementary Methods and Supplementary Results. It took 5-8 minutes to fill out the self-report questionnaire when tested on 10 co-authors.

**Table 2:**
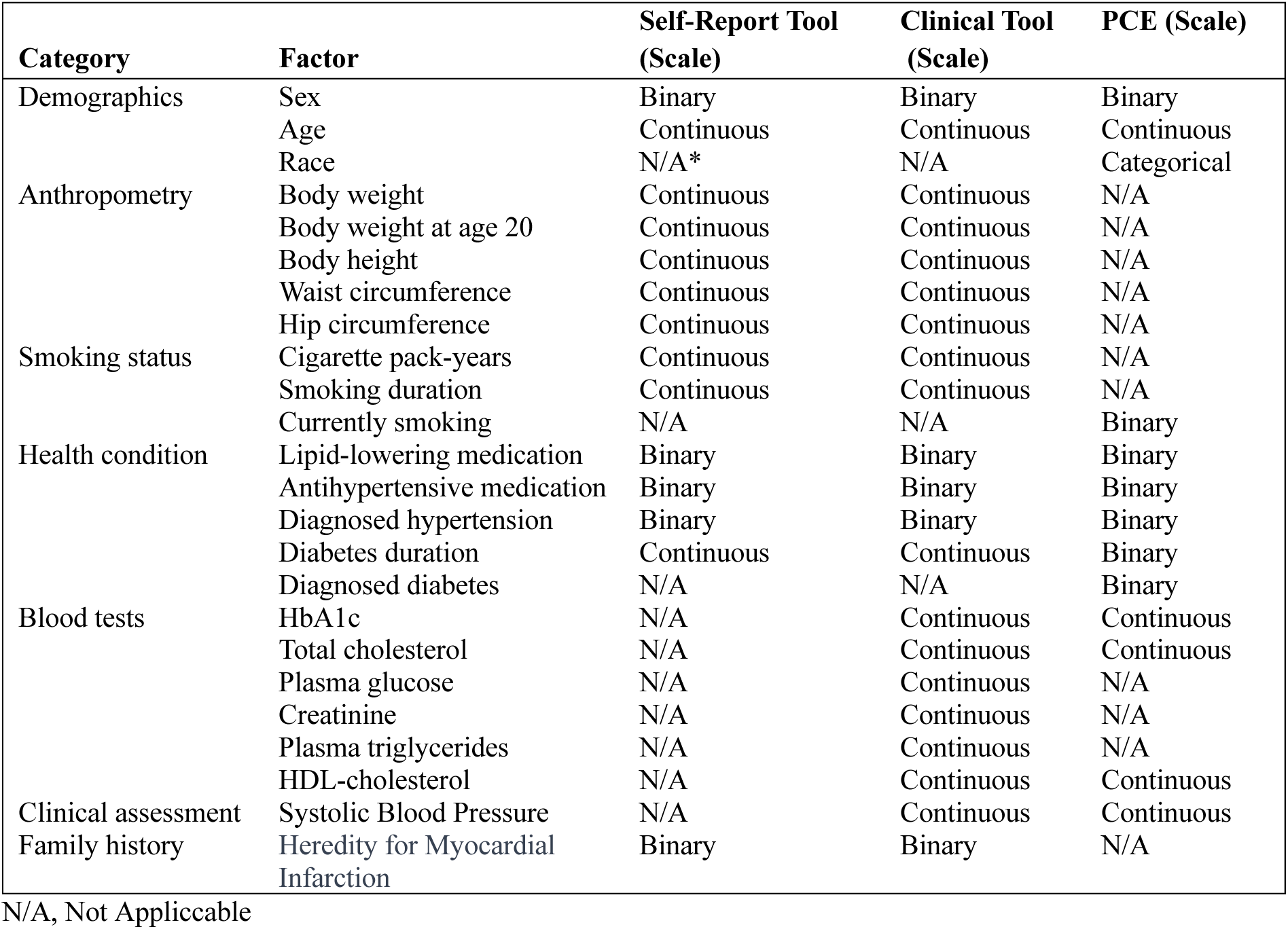
Factors in the self-report, clinical tool and the Pooled Cohort Equation (PCE) and their measurement scales.

### Prediction tools

The self-report tool had a high-to-excellent discriminatory capacity for SIS ≥4 in the external validation cohort (AUC 0.79, 95% CI: 0.76-0.83); and its discriminatory capacity was significantly better than the PCE risk score (AUC 0.76, 95% CI: 0.75-0.78, p<0.005, Figure 1A). With respect to variable importance of the self-report tool, sex and age were the most important variables, followed by cigarette pack-years, lipid-lowering medication, smoking duration, and several anthropometric measures (Figure 1B). The clinical tool performed slightly better than the self-report tool (AUC 0.80, 95% CI: 0.77-0.84, p<0.05, Figure 1C).

**Figure 1:**
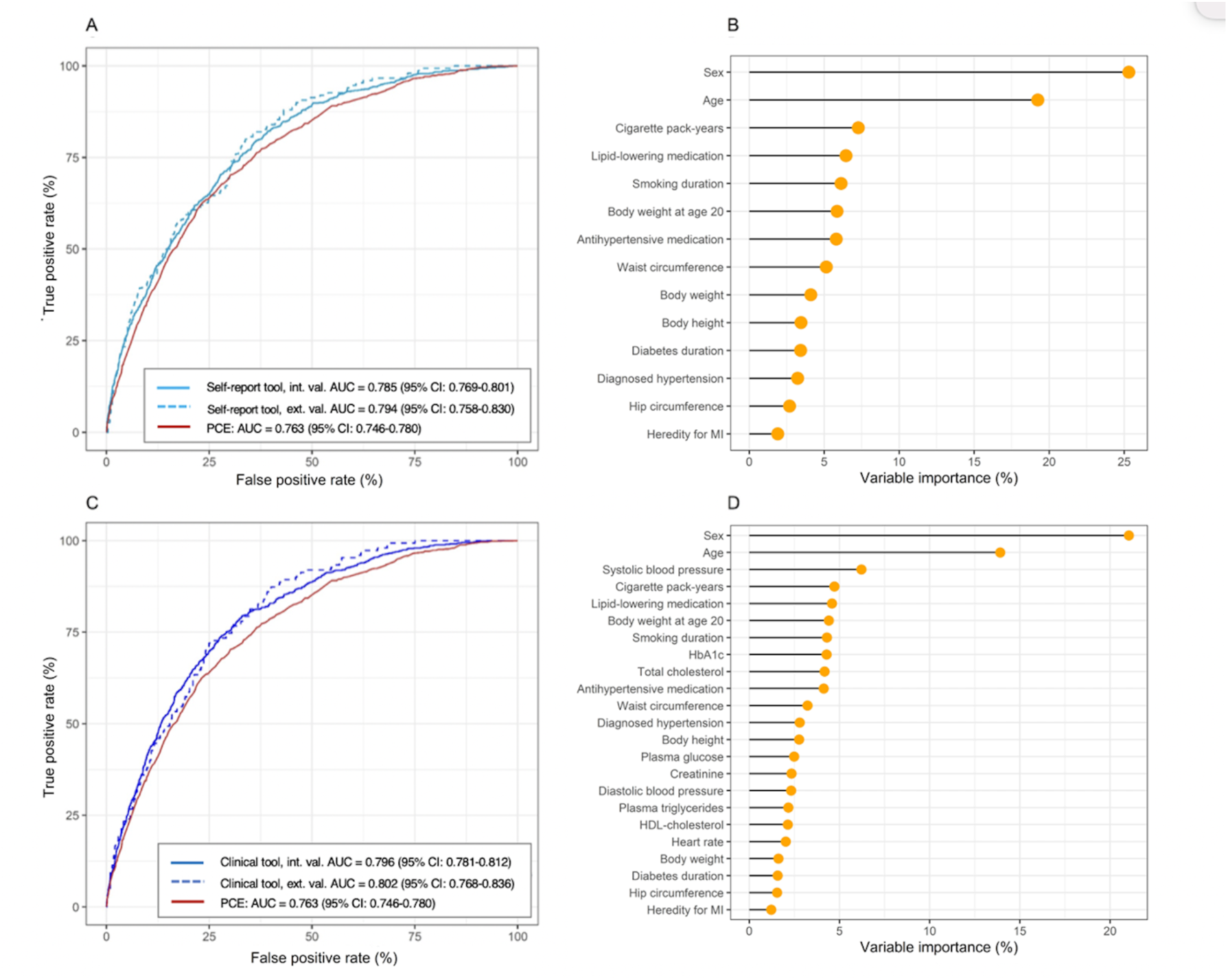
Receiver operating characteristic (ROC) curve for the self-report tool’s assessment of SIS≥4 in the internal and external validation group compared to PCE (Panel A, self-report tool vs. PCE, p <0.001, p <0.001 for internal and external validation respectively). Variable importance of the self-report tool (Panel B). ROC curve for the clinical tool’s assessment of SIS≥4 compared to PCE (Panel C, clinical tool vs. PCE, p <0.001, p <0.001 for internal and external validation respectively). Variable importance of the clinical tool (Panel D). AUC, area under the curve. The DeLong test was used for statistical comparison.

The variable importance of the clinical tool is presented in Figure 1 D, where systolic blood pressure and laboratory measurements of HbA1c and total cholesterol also were important for prediction. Both tools were well-calibrated (Supplementary Figure S3). SHAP plots for the interpretation of each variable’s contribution to the tools are presented in Supplementary Figures S4-S5. Results for the prediction of CACS ≥100 were largely similar (Supplementary Figure S6-S9).

### Sensitivity and stratification analyses

In general, both prediction tools performed better in women than in men (Figure 2); and they performed better in individuals older than 55 years of age (Figure 3). Similar results were seen for CACS (Figure S10 and S11). The two tools were largely unaffected by stratification for education, country of birth, and financial resources (Supplementary Table S5 and S6). As an example, the AUC of the self-report tool was 0.79 (95% CI: 0.76-0.81) in university educated and 0.78 (95% CI: 0.76-0.78) in non-university educated. The tool that was developed based on all 127 factors had an AUC of 0.80 (95% CI: 0.78-0.81).

**Figure 2:**
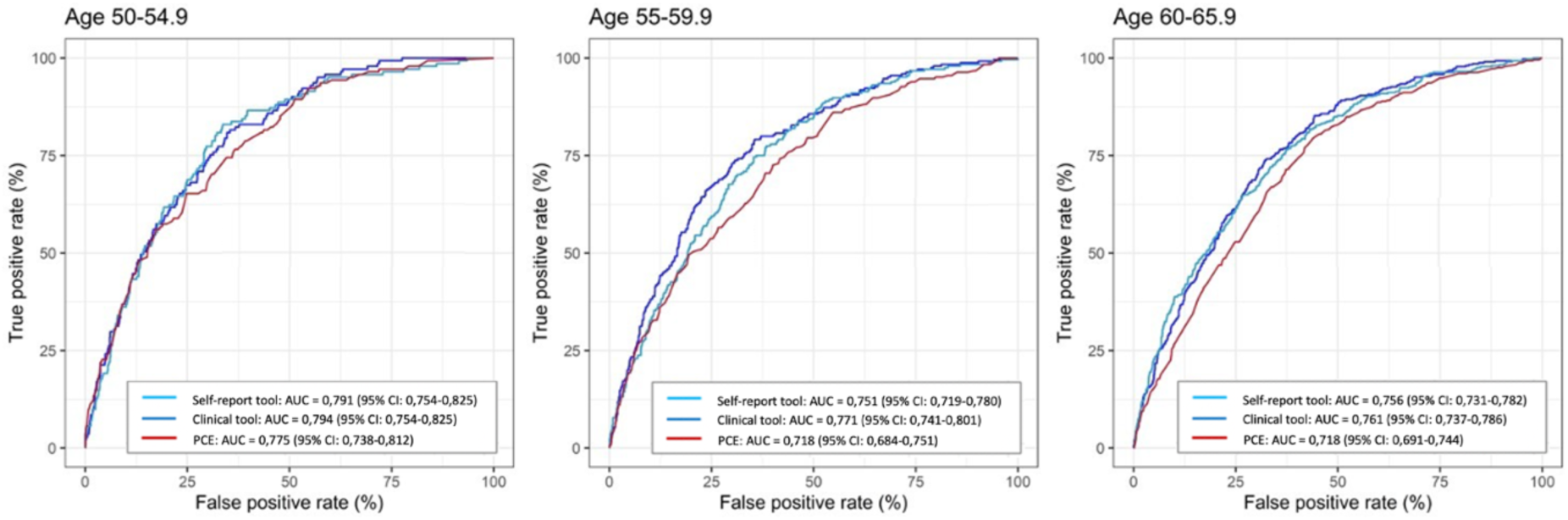
Receiver operating characteristic curves for the self-report tool and the clinical tool assessing SIS ≥4 vs. PCE, stratified by age. Age 50-54.9 (NS, NS for self-report and clinical tool respectively), 55-59.9 (p <0.05, p <0.001 for self-report and clinical tool respectively), 60–65.9 (p <0.001, p <0.001 for self-report and clinical tool respectively). The DeLong test was used for statistical comparison. NS; non-significant.

**Figure 3:**
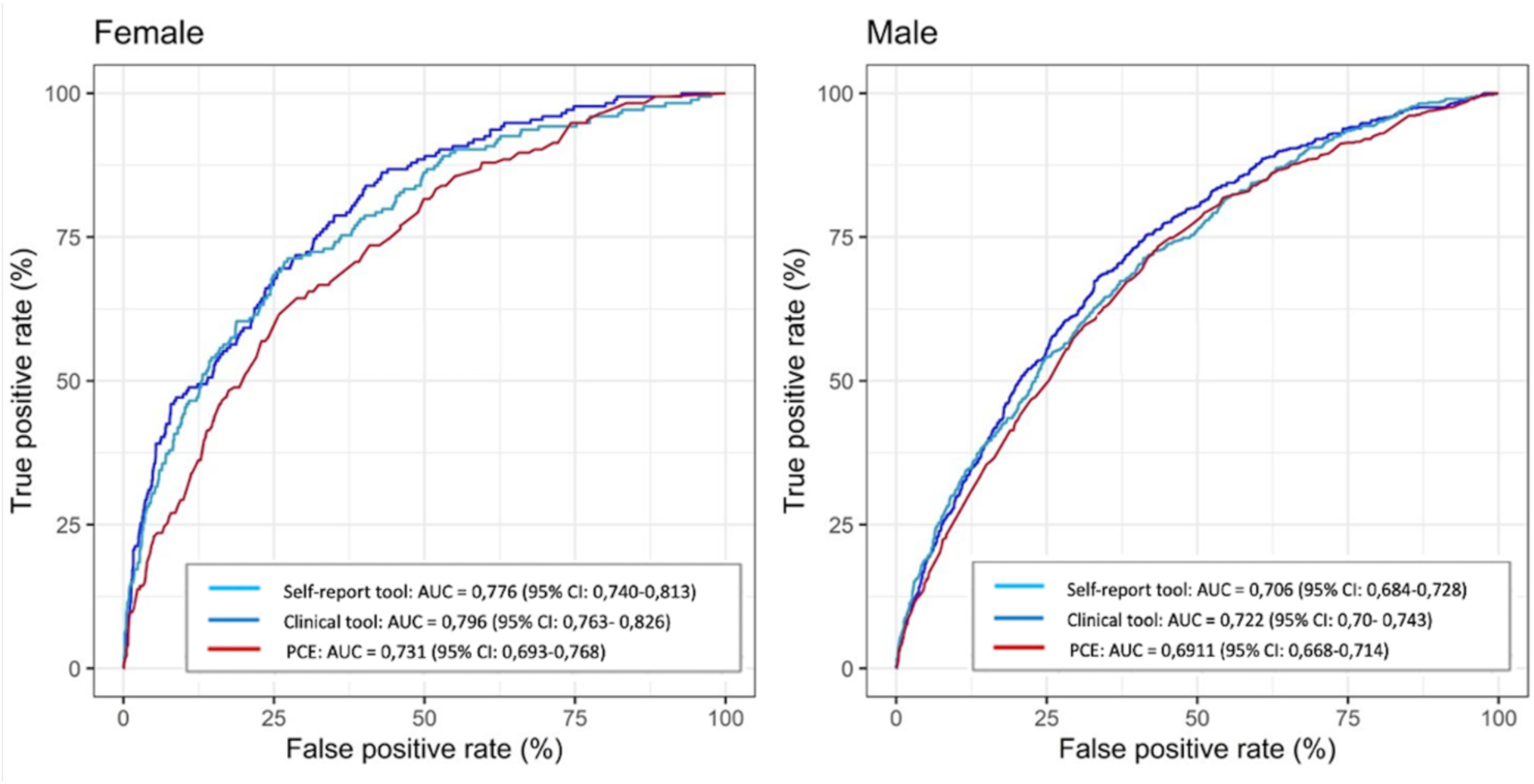
Receiver operating characteristic curves for the self-report tool and the clinical tool assessing SIS ≥4 vs. PCE, stratified by sex. Female; Self-report tool vs. PCE (p < 0.001), Clinical tool vs. PCE (p < 0.001). Male; Self-report tool vs. PCE (p < 0.001), Clinical tool vs. PCE (NS). The DeLong test was used for statistical comparison. NS; non-significant.

### Population-ordered distribution and decision curves

To exemplify how the prediction tools can be applied in a screening situation, we divided the population into ten groups of equal size, ordered by predicted risk (Table 3). Upon visual inspection, we identified a high-risk group in the individuals with the top 30% of assessed risk (i.e. top three deciles), with a mean absolute risk of SIS ≥4 of 27.1% for the self-report tool as indicated by red markings in Table 3. The middle four deciles suggested a moderate mean absolute risk, averaging 9,3% and marked with yellow. Additionally, a low-risk group was identified in the individuals in the bottom 30% of assessed risk, with a mean absolute risk of SIS ≥4 of 2.4%, marked green. The clinical tool showed similar patterns (Supplementary Table 7).

**Table 3:** Population-ordered distribution for the self-report tool in identifying Segment Involvement Score (SIS) ≥4. The first decile presents individuals at the highest mean absolute risk. Red – High mean absolute risk (average risk 27.1%); Yellow – Moderate mean absolute risk (average risk 9.3%); Green – Low mean absolute risk (average risk 2.4%).

| Decile of risk | Cumulative % of population | Mean absolute risk for SIS$\geq 4$ (%) | Cumulative number of CCTAs per finding | Cumulative % of all positives | Age (years)* | Male (%) | SIS (number of segments)* | CACS (Agatston Score)* | BMI (kg/m<sup>2</sup>)* |
| --- | --- | --- | --- | --- | --- | --- | --- | --- | --- |
| 1 | 10% | 39.5 | 2.5 | 31.4 | 61.9 | 92.3 | 3.1 | 240.2 | 28.6 |
| 2 | 20% | 23.0 | 3.2 | 49.7 | 61.0 | 91.1 | 2.0 | 92.1 | 27.7 |
| 3 | 30% | 18.7 | 3.7 | 64.6 | 59.0 | 81.1 | 1.8 | 87.7 | 27.7 |
| 4 | 40% | 11.4 | 4.3 | 73.7 | 57.4 | 70.6 | 1.3 | 55.2 | 27.9 |
| 5 | 50% | 12.4 | 4.8 | 83.6 | 57.3 | 55.7 | 1.2 | 40.6 | 27.0 |
| 6 | 60% | 7.1 | 5.3 | 89.3 | 57.4 | 43.5 | 0.8 | 28.3 | 26.8 |
| 7 | 70% | 6.2 | 5.9 | 94.2 | 57.1 | 38.1 | 0.7 | 20.9 | 25.8 |
| 8 | 80% | 3.7 | 6.6 | 97.1 | 56.2 | 21.9 | 0.6 | 15.5 | 26.0 |
| 9 | 90% | 2.4 | 7.2 | 99.0 | 54.3 | 6.8 | 0.4 | 12.0 | 25.9 |
| 10 | 100% | 1.3 | 8.0 | 100.0 | 52.5 | 0.8 | 0.2 | 3.9 | 24.8 |

SIS, Segment Involvement Score; CCTA, Coronary Computed Tomography Angiography; CACS, Coronary Artery Calcium Score; BMI, Body Mass Index. \* mean.

The remaining 40% constituted a moderate risk group, with a mean absolute risk of SIS ≥4 of 9.3% and 8.8% for the self-report and clinical tool, respectively (marked yellow in the tables). Among individuals who fell within the high-risk group, we identified 64.6% and 67.3% of all individuals with SIS ≥4 using the self-report and clinical tool, respectively. In order to find one individual with SIS ≥4 in the high-risk group, an average of 3.7 CCTAs have to be performed for the self-report tool and 3.5 CCTAs for the clinical tool. Similar results were observed for CACS ≥100 (Supplementary Table S8 and S9).

Decision curve analyses for SIS ≥4 and CACS ≥100 showed that both the self-report and the clinical tool is superior to the PCE when applied to a population with a mean absolute risk corresponding to the high-risk group (i.e. 27-28%) (Supplementary Figure S11 and S12).

## Discussion

This study shows that we can effectively use non-imaging data to identify individuals from the general population with a high likelihood of having moderate to severe coronary atherosclerosis. The discriminative ability to identify SIS ≥4 and CACS ≥100 was high-to-excellent in the external validation cohort (AUC 0.79-0.81). Most importantly, the self-report tool, based only on data that do not require a health care visit, performed almost equal to the clinical tool, which included blood tests and physical examinations.

These results suggest that screening for coronary atherosclerosis can be performed with only self-reported data and that the screening is only marginally improved by a visit to a healthcare facility. This corroborates earlier findings, in which other self-report tools for cardiovascular risk prediction have shown promising results compared to traditional clinical risk assessment tools for the 10-year risk of cardiovascular death.^11,12^ In addition, three main risk groups were identified in our analyses of the population ordered distribution of risk: a high-risk group, a moderate-risk group and a low-risk group. In the high-risk group (top 30% of the mean absolute risk), up to 65% of all individuals with moderate to severe coronary atherosclerosis can be identified via the self-report tool (65% of those with SIS ≥4 and 64% of those with CACS ≥100). In contrast, only 3-4% of all individuals with moderate to severe coronary atherosclerosis is found in the low-risk group (bottom 30% of the mean absolute risk).

If our results are recalculated as the number needed to screen, a total of 3.7 CCTA examinations must be performed among high-risk individuals to detect one individual with SIS ≥4 using the self-report tool (3.5 CCTA for the clinical tool). Further, the proportion of individuals with SIS ≥4 would be around 27 to 28% in the same high-risk group. Our decision curve analysis showed that both the self-report and the clinical tool were superior to the PCE at this threshold probability. Results for CACS were similar.

Implementation of a strategy that combines self-reported risk and subsequent imaging for SIS or CACS would result in the targeted identification of individuals who would benefit from improved lipid-lowering or other preventive therapies^1,2^. Future studies will have to test the cost-effectiveness of such a screening strategy to reduce the overall burden of cardiovascular disease.

We used a combination of data-driven and manual factor selection to optimize the performance of our tools. The strategy focused on the most easily accessible factors with the greatest impact on model performance. Not surprisingly, information on traditional risk factors^1^, such as sex, age, lipid-lowering medication, systolic blood pressure, anti-hypertensive medication, diabetes, and smoking status, were influential for both the self-report and the clinical tool. In addition, our data selection and variable importance emphasized the significance of continuous data such as smoking duration and cigarette pack-years, rather than binary factors such as current smoking, yes/no. It is also noteworthy that many anthropometric measures ranked high in variable importance, confirming the importance of body weight^25^ and abdominal obesity.^26^ Weight at age 20, previously linked to an elevated risk of coronary atherosclerosis ^27^, was one of the most important non-traditional factors, surpassing the impact of weight at the time of examination. On the other hand, a parent’s history of myocardial infarction before age 60, ranked low in variable importance.

Family history of myocardial infarction is included in some^28,29^, but not all^1^, of the previous clinical risk algorithms. We were reassured that the data reduction was successful, given that a tool that utilizing all 127 available factors achieved an AUC similar to the clinical tool.

Using data from the LifeLines imaging trial (ImaLife), Ties and co-workers^30^ tested a pre-screening strategy to identify individuals with a high probability of having CACS ≥100. They showed that using a risk algorithm (SCORE2), primarily developed to identify the 10-year risk of developing a cardiovascular event, was not very efficient to identify individuals with CACS ≥100. If one or more traditional risk factors were present, around 89% of all individuals with CACS ≥100 were identified; however, using that strategy, around 70% of the population had to be imaged. Our combination of self-reported risk factors into a risk assessment tool appears to be slightly more efficient, as we could identify 96% of all individuals with CACS ≥100 with the same percentage of population imaging.

### Limitations

A number of limitations of the current study must be acknowledged. First, all population-based studies suffer from recruitment bias, often resulting in a lower cardiovascular risk level compared to the background population. This is also true in SCAPIS^20,31^ and, therefore, we tested whether recruitment bias could have affected the results by stratifying our sample by several socio-economic factors. However, our tools performed equally well in the different subgroups, suggesting that recruitment bias does not affect the performance of the tools to a major extent. Second, the derivation and validation populations were mainly of northern European descent and the results may not be generalizable to other geographical regions.

Third, we mainly used traditional risk factors as predictors in our tools. It is possible that non-traditional risk factors derived from further biochemical or genetic analyses could aid prediction beyond what was presented here. Fourth, our self-report tool was based on data acquired at the test sites in SCAPIS and not from home. Truly self-reported data may introduce more inaccurate measurements and lower the performance of the tools. Fifth, we chose SIS and CACS as our outcomes since they are robust and easily accessible measures of coronary atherosclerosis. In the future, we may identify other phenotypes of coronary atherosclerosis that confer a higher risk of cardiovascular disease. Sixth, our external validation relied on a pilot trial to the main study. Although using unique individuals and a fundamentally different study protocol, this may impact the generalizability of our findings to broader populations.

### Clinical implications

The presence of coronary atherosclerosis confers an increased risk of future myocardial infarction. Here, we present a self-report tool, trained and validated in populations without history of previous IHD, which effectively identified individuals with moderate to severe coronary atherosclerosis using self-reported data alone. Our results on the self-reported data open up for the possibility to develop cost-effective screening programs and extend previous observations on self-reported data to identify high-risk individuals for clinical IHD.^11,12^ The self-report tool can be used on its own to identify high-risk individuals or to select individuals in need of further risk refinement via imagining. It is also possible that this tool can be effective in the communication with individuals about behavioral changes, given that sharing of medical imaging information has been shown to affect smoking, eating, and physical activity behaviors.^32^

## Conclusion

We have developed a prediction tool based only on self-reported data that with good-to-excellent discriminative ability can identify individuals with moderate to severe coronary atherosclerosis. The tool performed equally well to a model also including clinical data generated after a clinical visit. The self-report tool could be the starting point for a screening program to identify high-risk individuals in need of imaging and further risk evaluation.

## Data Availability

Due to the nature of the sensitive personal data and study materials they cannot be made freely available. However, by contacting the corresponding author or study organization (www.scapis.org), procedures for sharing data, analytic methods, and study materials for reproducing the results or replicating the procedure can be arranged following Swedish legislation.

## Acknowledgement

We are grateful to all the participants in SCAPIS and would like to give a special thanks to all test personnel at the SCAPIS test centers around Sweden.

